# FACTORS AFFECTING MENSTRUAL HYGIENE AMONG WOMEN OF SLUM AREA OF BUDANILKANTHA MUNICIPALITY, KATHMANDU

**DOI:** 10.1101/2025.10.26.25338843

**Authors:** Bijaya Laxmi Niraula, Akshaya Acharya, Manoj Phuyal, Baburam Marasini, Puspa Acharya

## Abstract

**Background:** Menstruation is often stigmatized and poorly managed in low-income settings like Nepal, leading to health and social challenges. Women in urban slums face additional barriers due to poverty and limited sanitation. This study explores factors affecting menstrual hygiene practices among women in Budanilkantha slums to inform targeted interventions.

**Methods:** A cross-sectional study was conducted among 247 women aged 18–49 years in slum areas of Budanilkantha Municipality, Nepal. Participants were selected using systematic random sampling. Data were collected via a pretested questionnaire and analysed with chi-square tests. Ethical approval was obtained from Nobel College, Pokhara University.

**Results:** Among the 247 women surveyed, 53.8% used sanitary pads while 42.9% used cloth during menstruation. Most practiced handwashing with soap and water (66.8%) and cleaned external genitalia similarly (82.6%), but only 38.9% bathed daily during menstruation. Nearly all participants (95.1%) experienced menstrual restrictions, with 57.1% perceived as “impure.” Awareness of menstruation before menarche was reported by 65.6%, and higher education and income were significantly associated with better awareness and hygiene practices.

**Conclusions:** Menstrual hygiene practices in Budanilkantha’s slum areas are influenced by socio-economic status, cultural taboos, and limited infrastructure. Although basic hygiene is practiced by many, cultural stigma and gaps in hygiene behaviours persist. Comprehensive interventions addressing education, WASH infrastructure, and social norms are essential to improve menstrual health and dignity among marginalized women.

## Introduction

Menstruation is a natural biological process marking the transition from childhood to womanhood. Despite its physiological normalcy, menstruation remains a subject of silence, stigma, and discrimination in many societies, particularly in low- and middle-income countries (LMICs) such as Nepal. Inadequate menstrual hygiene management (MHM) has been associated with negative health outcomes, including reproductive tract infections, urinary tract infections, and psychological distress. Moreover, it affects educational attainment, mobility, and the overall well-being of adolescent girls and women [1–2].

Globally, more than 500 million women and girls are estimated to lack access to adequate menstrual hygiene facilities and products [3]. The challenge is even greater in urban slums where poverty, poor sanitation, limited water supply, and deep-rooted cultural taboos hinder proper menstrual hygiene practices [4]. In Nepal, although national efforts have been made to address adolescent health, the management of menstruation remains under-recognized in public health policy. A national survey conducted in 2019 reported that while 71.7% of adolescent girls knew about menstruation before menarche, nearly half lacked understanding of its physiology, and many experienced social restrictions and stigma during their menstrual cycles [5]

Women and girls residing in urban slum areas, such as those in Budanilkantha Municipality in Kathmandu, often face compounded vulnerabilities due to their socio-economic conditions, inadequate sanitation infrastructure, and lack of access to health education. These factors collectively impact their ability to manage menstruation safely and with dignity [6]. Various studies have identified socio-demographic characteristics including age, education, religion, maternal literacy, and income as significant determinants of menstrual hygiene practices [7–9].

Despite existing evidence from different parts of Nepal and South Asia, there is a dearth of localized data focusing on menstrual hygiene among women living in slum areas of Kathmandu Valley. This study was, therefore, conducted to identify the factors influencing menstrual hygiene practices among women aged 18–45 residing in slum communities of Budanilkantha Municipality. Understanding these factors is essential for informing targeted interventions and policy measures aimed at improving menstrual health outcomes and ensuring gender equity in hygiene and sanitation.

## Materials and Methods

### Methods

This community-based analytical cross-sectional study aimed to assess menstrual hygiene practices and associated factors among women living in selected slum areas of Budanilkantha Municipality, Kathmandu, Nepal. The study was conducted from July 25 to November 2022 in a semi-urban setting characterized by informal settlements with limited access to water, sanitation, and hygiene (WASH) services.

### Study Participants

The study population consisted of women aged 18–49 years residing in slum areas of Budanilkantha Municipality. Women were eligible for inclusion if they had lived in the selected slum areas for at least six months, were willing to participate, and provided written informed consent. In cases where the head of the household was unavailable during data collection, another responsible adult family member was consulted to facilitate access. Women were excluded if they did not reside in the designated slum areas, were unable to communicate due to serious health conditions, or declined to provide consent.

### Sampling Technique and Sample Size

The sample size was calculated using the single population proportion formula:

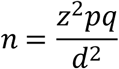

The sample size was calculated using the formula for estimating a proportion in a finite population, assuming a 39% prevalence of good menstrual hygiene practices [10], a 5% margin of error, and a 95% confidence interval. After accounting for a 10% non-response rate, the final sample size was 247.

The study was conducted in four wards of Budanilkantha Municipality (Wards 4, 6, 8, and 10), which contain slum settlements. These wards were purposively selected as they represent all slum areas in the municipality. A total of 1,145 slum households were listed in collaboration with local health authorities, with 578 eligible women aged 18–49 years identified. Using the Probability Proportionate to Size (PPS) technique, a sample size of 247 women was proportionally allocated to each ward. Based on the number of eligible women in each ward, 25 respondents were selected out of 58 in Ward 4, 53 out of 123 in Ward 6, 153 out of 357 in Ward 8, and 16 out of 40 in Ward 10. Households were selected using systematic random sampling. The first household was chosen based on the bottle spin method, and if no eligible woman was found, the next consecutive household was selected. One eligible woman per household was interviewed.

### Data Collection Procedures

Data were collected using a pre-tested, semi-structured questionnaire developed based on a comprehensive review of relevant literature. The questionnaire was initially prepared in English, then translated into Nepali and back-translated to ensure accuracy and consistency. Key sections of the tool covered socio-demographic characteristics, knowledge and practices related to menstrual hygiene, access to WASH (Water, Sanitation, and Hygiene) facilities, and sociocultural restrictions surrounding menstruation. A pilot test was conducted among 10% of the total sample size in a similar slum area not included in the main study, and necessary adjustments were made based on the feedback received. Data collection was carried out through face-to-face interviews by trained enumerators. Field supervisors and the principal investigator performed daily reviews to ensure completeness and accuracy of the collected data.

### Variables

Independent variables included age, marital status, education, occupation, family income, ethnicity, religion, family structure, knowledge of menstruation, and cultural restrictions. The outcome variable was menstrual hygiene practice, categorized as “appropriate” or “inappropriate” based on definitions established in the literature.

### Statistical Analysis

Data were entered into EpiData (version 3.1) and analyzed using SPSS (version 16). Descriptive statistics (means, standard deviations, frequencies, percentages) were used to summarize data. Associations between menstrual hygiene practices and independent variables were examined using Chi-square tests. A p-value < 0.05 was considered statistically significant. Missing data and incomplete questionnaires were reviewed and excluded from statistical analysis where necessary.

### Ethical Statement

Ethical approval for this study was obtained from the Institutional Review Committee of Nobel College, Pokhara University (Reference no. 01). Written informed consent was obtained from all participants prior to inclusion in the study; for non-literate participants, thumbprints were accepted as consent. Participants were informed of their right to withdraw at any time without penalty. Confidentiality and anonymity of participant information were maintained by using anonymized codes and secure storage of data in password-protected computers and locked cabinets. This study was conducted in accordance with the ethical principles of the Declaration of Helsinki (2013 revision).

## Results

### Socio-Demographic and Road Traffic Accident (RTA) Characteristics

Table 1 summarizes the socio-demographic characteristics and menstrual hygiene practices of the 247 women included in the study. The majority of respondents (48.2%) were aged between 20 and 25 years, with a mean age of 24.8 years. Most participants were Hindu (69.7%), followed by Buddhists (27.9%). A significant proportion of respondents had only completed primary (34.4%) or secondary (25.1%) education, while just 5.7% had completed a bachelor’s degree or higher.

**Table 1.**
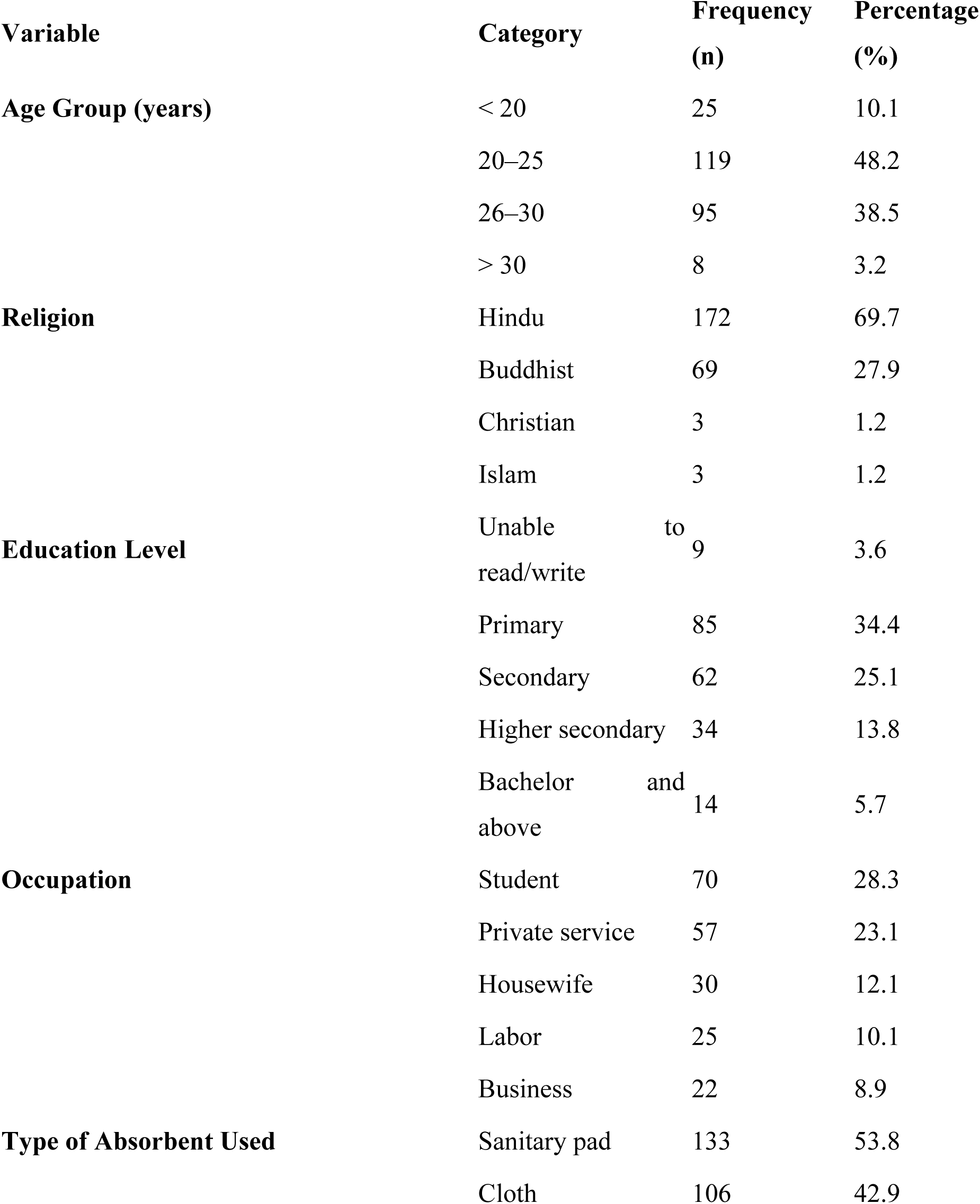
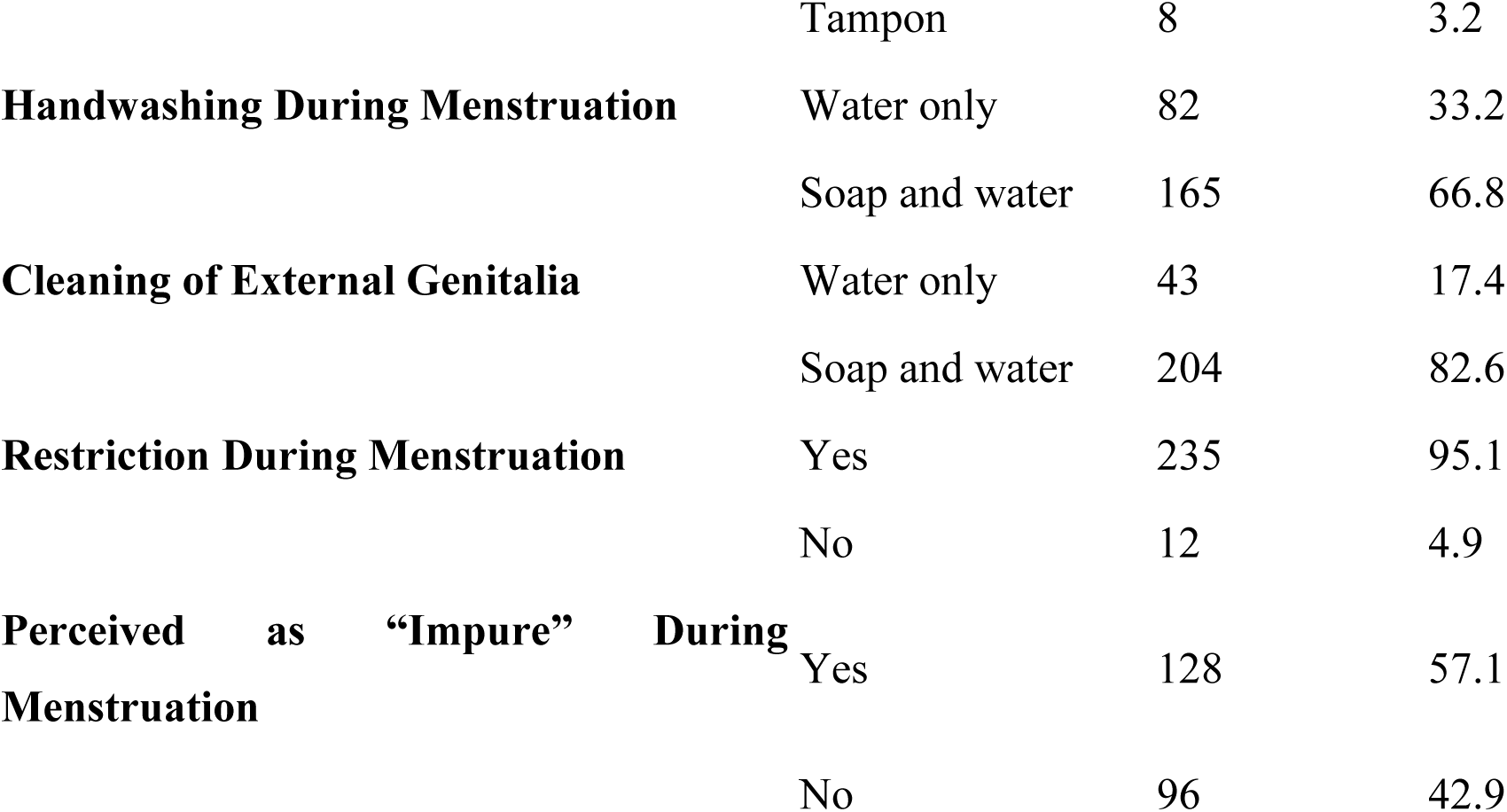
Socio-demographic Characteristics and Menstrual Hygiene Practices of Respondents.

In terms of occupation, students comprised the largest group (28.3%), followed by women working in private services (23.1%) and housewives (12.1%). Regarding menstrual hygiene practices, 53.8% of the participants reported using sanitary pads during menstruation, while 42.9% used cloth and 3.2% used tampons.

Hand hygiene during menstruation was generally good, with 66.8% of respondents using soap and water to wash their hands, although 33.2% relied on water only. Similarly, 82.6% of the women reported cleaning their external genitalia with soap and water, while 17.4% used only water.

Nearly all respondents (95.1%) reported facing some form of restriction during menstruation. Cultural stigma was evident, as more than half of the participants (57.1%) reported being perceived as "impure" during menstruation. These findings reflect not only the socio-economic challenges but also the deep-rooted cultural barriers that influence menstrual hygiene practices among women living in slum areas.

Table 2 presents the reinforcing sociocultural factors influencing menstrual hygiene practices among respondents. A substantial majority (95.1%) of women reported facing restrictions during menstruation. Among those who were restricted, the most common forms of restriction included household work (96.5%) and participation in celebrations (26.8%). Only a negligible number reported restrictions related to food (0.4%) and prayer (0.4%).

**Table 2.**
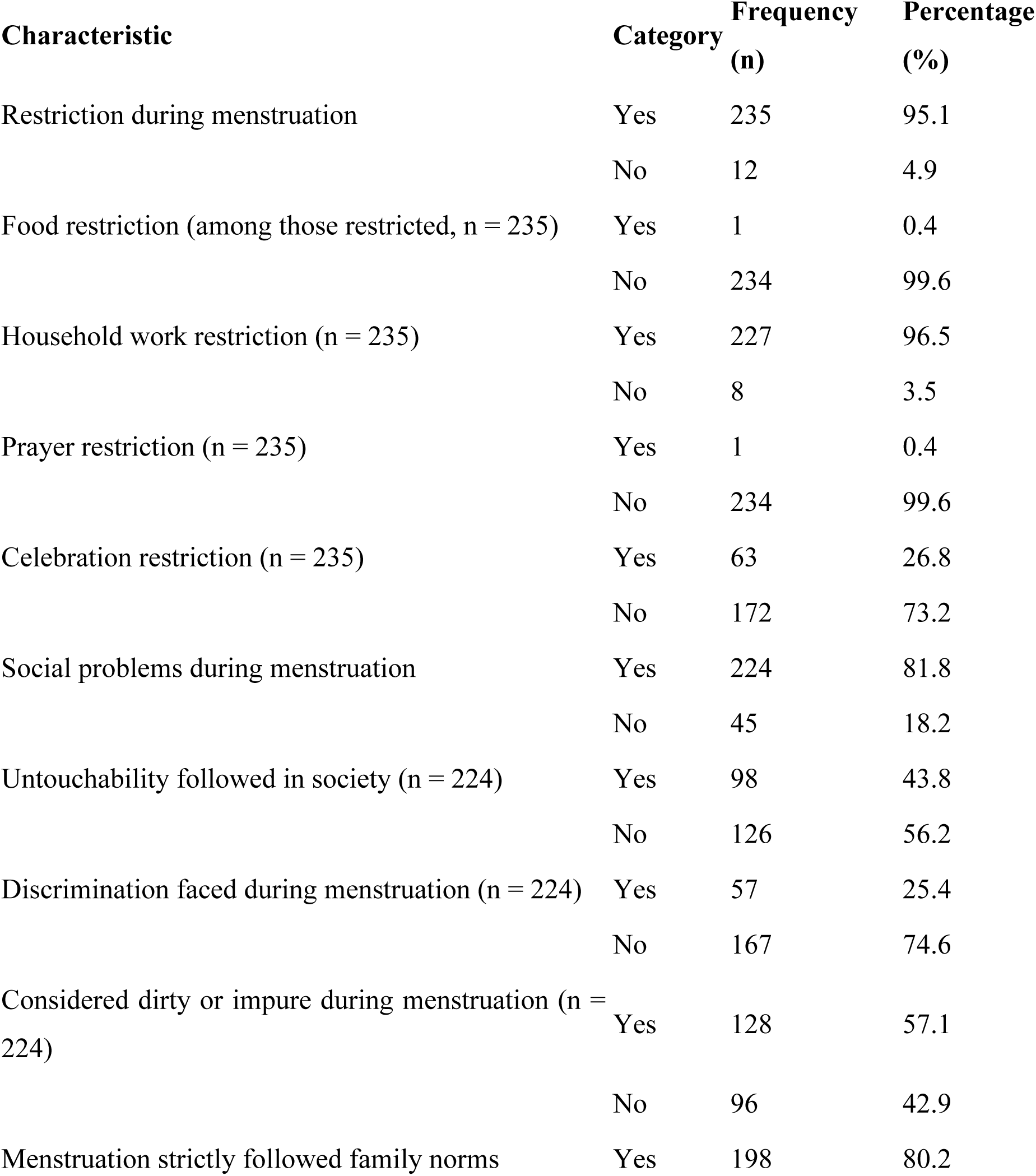
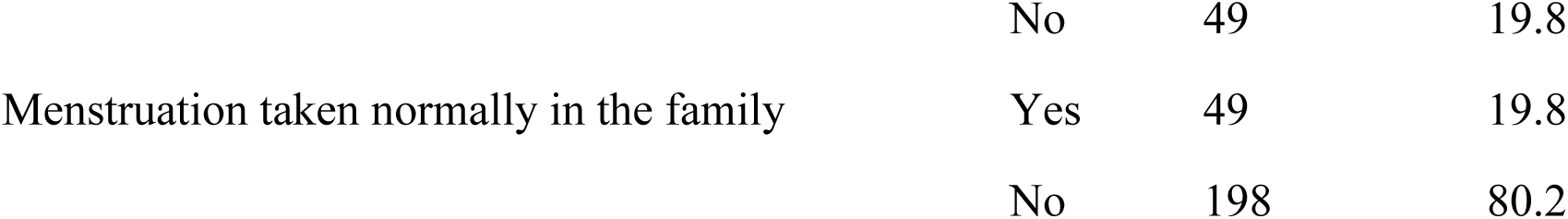
Reinforcing Factors Related to Menstrual Hygiene Among Respondents.

Social stigma surrounding menstruation was prevalent. More than four out of five respondents (81.8%) reported facing social problems during menstruation, and nearly half (43.8%) reported being subjected to untouchability practices. About one-quarter (25.4%) experienced direct discrimination during their menstrual cycle. Furthermore, 57.1% of participants reported being perceived as dirty or impure during menstruation.

In terms of family beliefs, 80.2% of women said that menstruation-related norms were strictly followed in their households, indicating the deep-rooted influence of traditional customs. Interestingly, only 19.8% of participants said menstruation was taken normally within their families, further underscoring the stigma and silence surrounding the topic.

Table 3 outlines the enabling factors and menstrual hygiene practices of the respondents. About two-thirds (65.6%) of women were aware of menstruation before its onset, and 88.7% reported receiving an adequate explanation when they first experienced it. The most common sources of menstrual information were family members (59.3%) and teachers (57.4%), followed by friends (30.7%) and social media (11.6%).

**Table 3.**
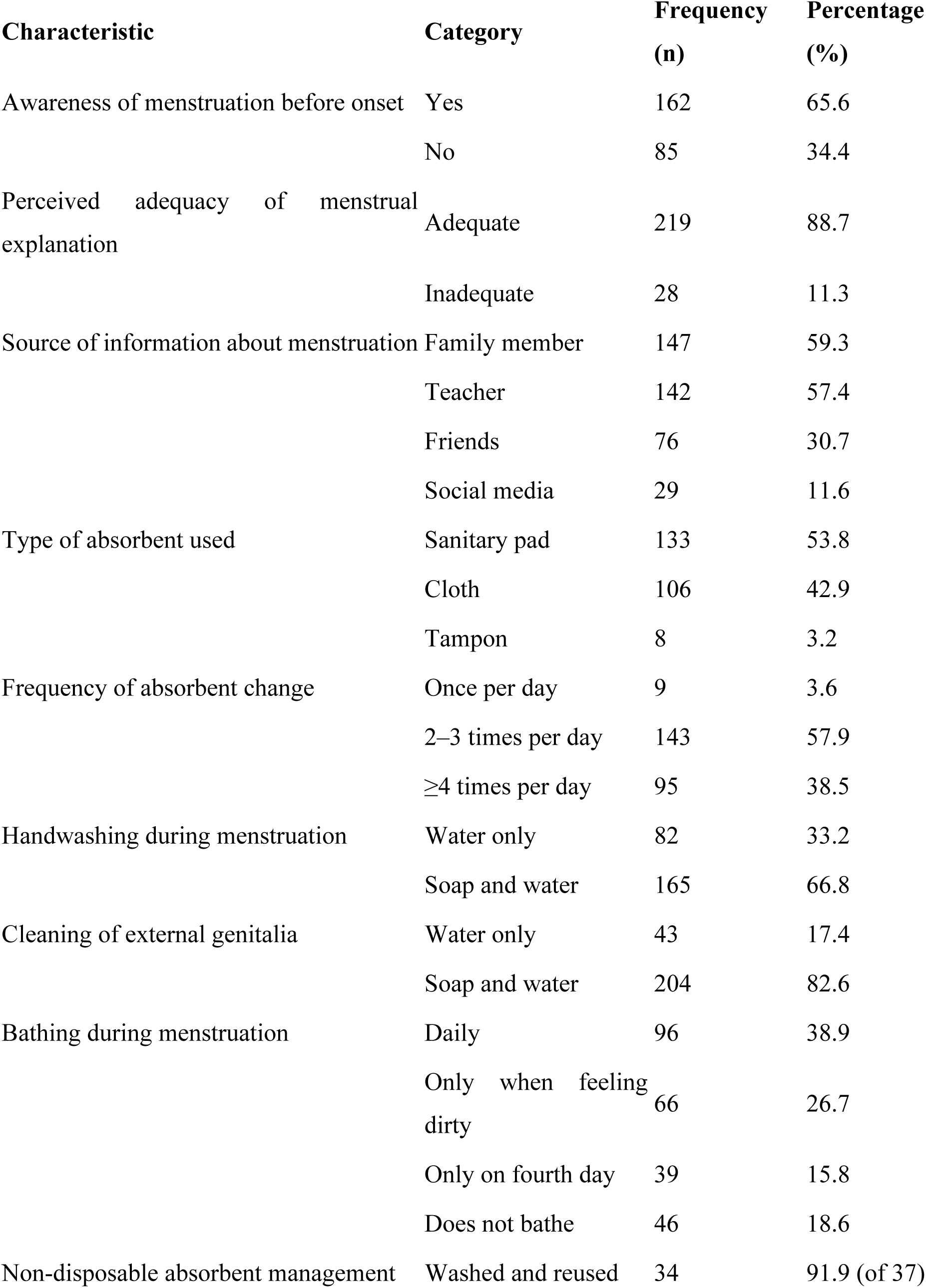
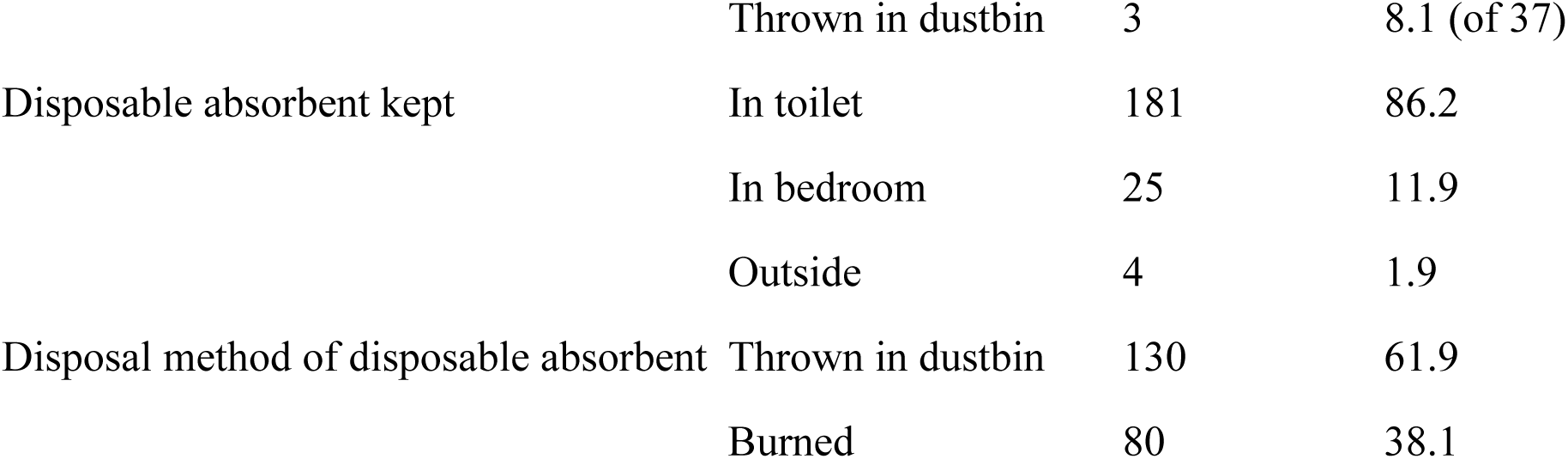
Enabling Factors and Menstrual Hygiene Practices Among Respondents.

In terms of menstrual hygiene materials, 53.8% of participants used disposable sanitary pads, while 42.9% used cloth, and 3.2% used tampons. The majority of women (57.9%) reported changing their absorbent materials two to three times per day, and 38.5% changed them four or more times per day. Only 3.6% changed absorbents once daily.

Regarding hygiene behaviors, 66.8% of women used soap and water for handwashing during menstruation, whereas 33.2% used only water. Similarly, 82.6% cleaned their external genitalia with soap and water, while 17.4% used only water.

Bathing frequency varied: 38.9% bathed daily during menstruation, 26.7% bathed when feeling dirty, 15.8% bathed only on the fourth day, and 18.6% did not bathe during menstruation. Among women who used non-disposable absorbents, the majority (91.9%) reported washing and reusing them, while a small proportion (8.1%) discarded them.

In terms of storing used disposable absorbents before disposal, 86.2% kept them in toilets, 11.9% in bedrooms, and 1.9% outside. For final disposal, 61.9% threw used absorbents into dustbins, while 38.1% burned them.

Table 4 shows the relationship between selected socio-demographic variables and awareness of menstruation before its onset among women in the study population. The analysis showed no statistically significant association between awareness and age (p = 0.858), with similar proportions of awareness observed across all age groups, ranging from 71.4% to 80.0%. Likewise, awareness did not vary significantly by religion (p = 0.600) or ethnicity (p = 0.938), indicating comparable levels of awareness among different religious and ethnic groups.

**Table 4.**
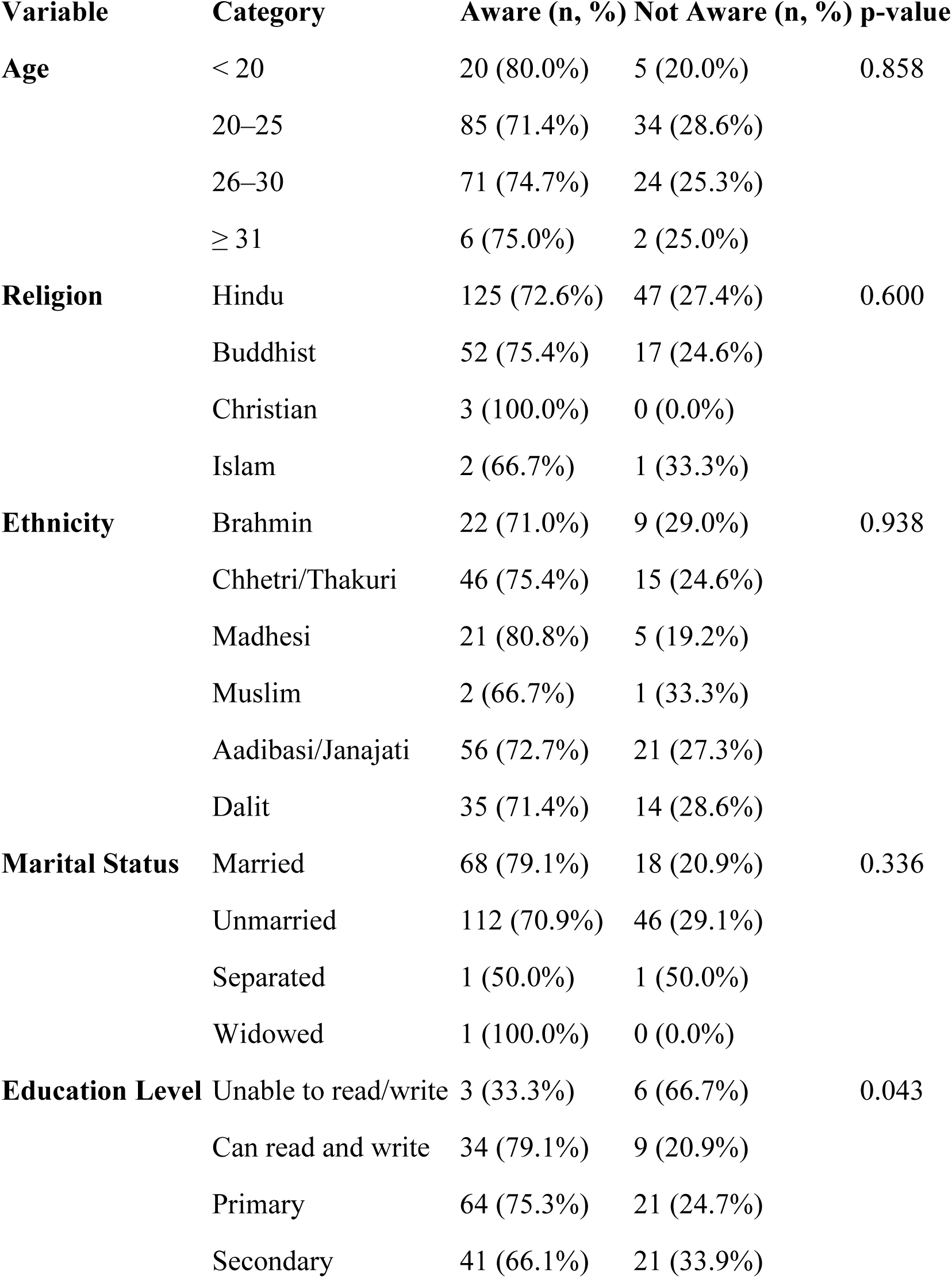
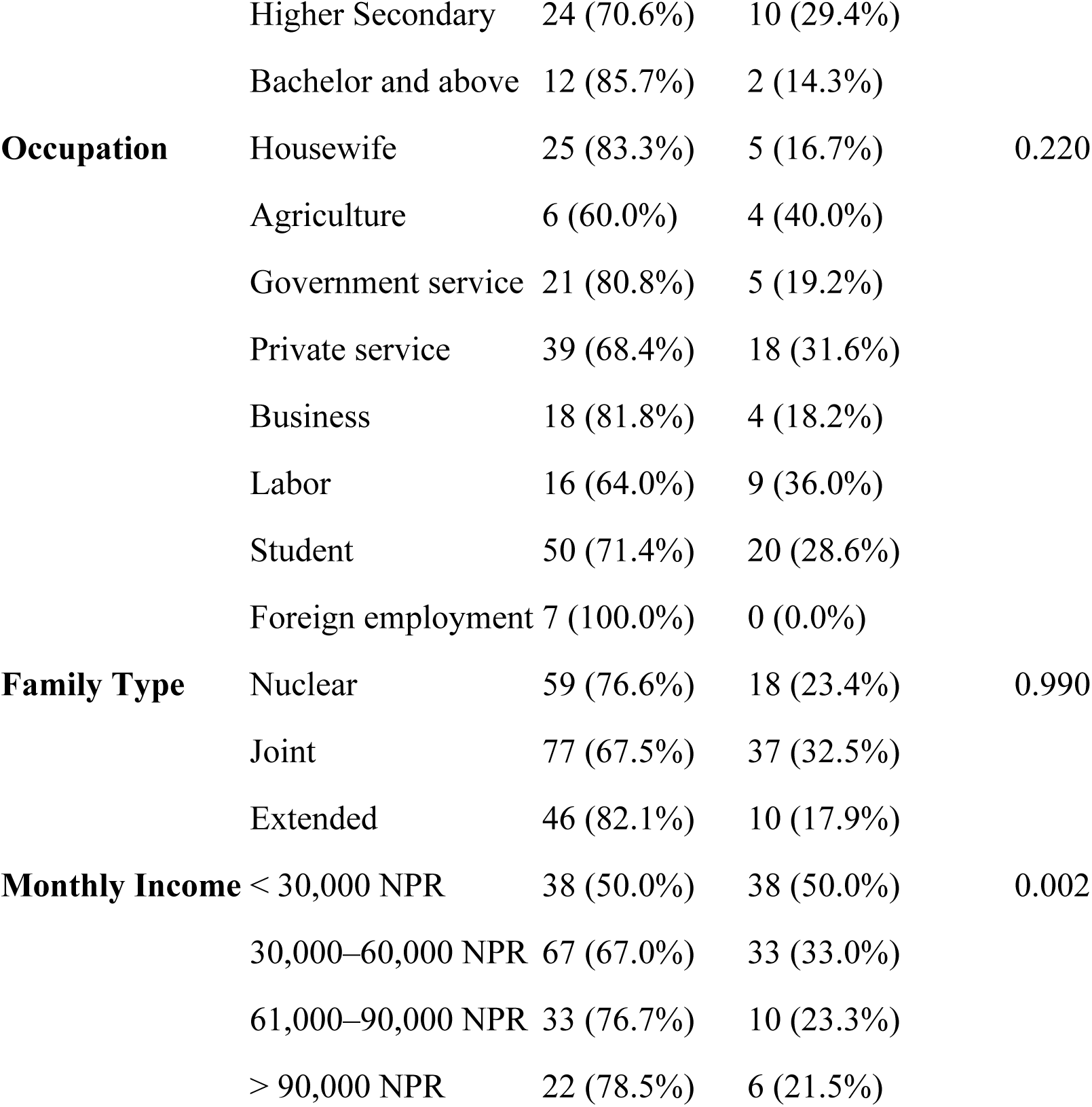
Association between Socio-demographic Characteristics and Awareness of Menstruation before Its Onset.

No significant difference was found between marital status and awareness (p = 0.336). However, awareness was slightly higher among married women (79.1%) compared to unmarried women (70.9%).

Education level was significantly associated with awareness (p = 0.043). Women unable to read and write had the lowest awareness (33.3%), while those with higher education, particularly women with a bachelor’s degree or above, exhibited the highest awareness levels (85.7%).

No significant relationship was observed between occupation and awareness (p = 0.220), although awareness varied across occupational groups, ranging from 60.0% among women involved in agriculture to 100% among those in foreign employment.

Family type (nuclear, joint, or extended) did not significantly influence awareness (p = 0.990). Lastly, monthly income was significantly associated with awareness (p = 0.002). Women with a monthly income below 30,000 NPR had the lowest awareness (50.0%), whereas awareness increased with income, reaching 78.5% among women earning more than 90,000 NPR per month.

Table 5 presents the relationship between socio-demographic characteristics and perceived adequacy of the explanation of menstruation.

**Table 5.**
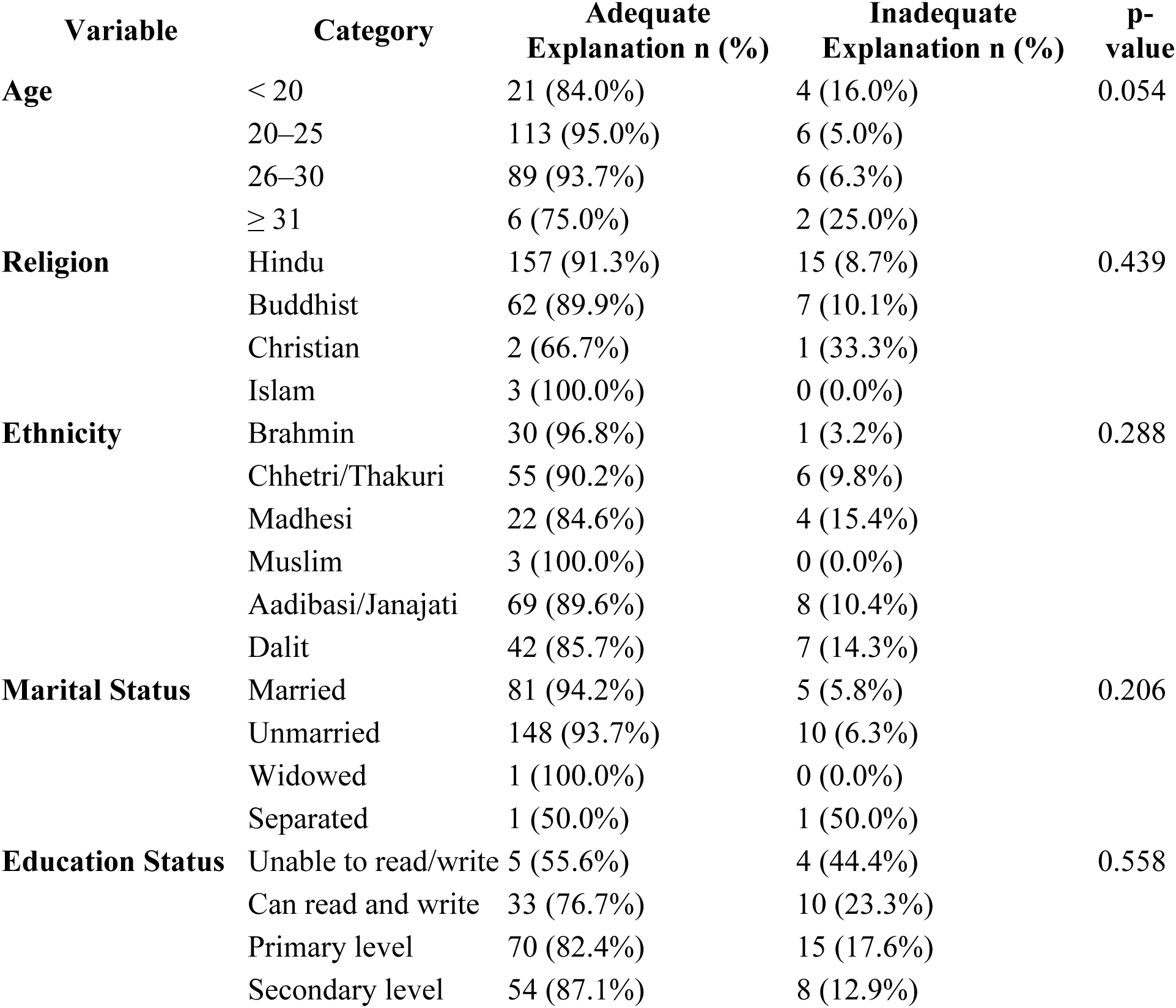
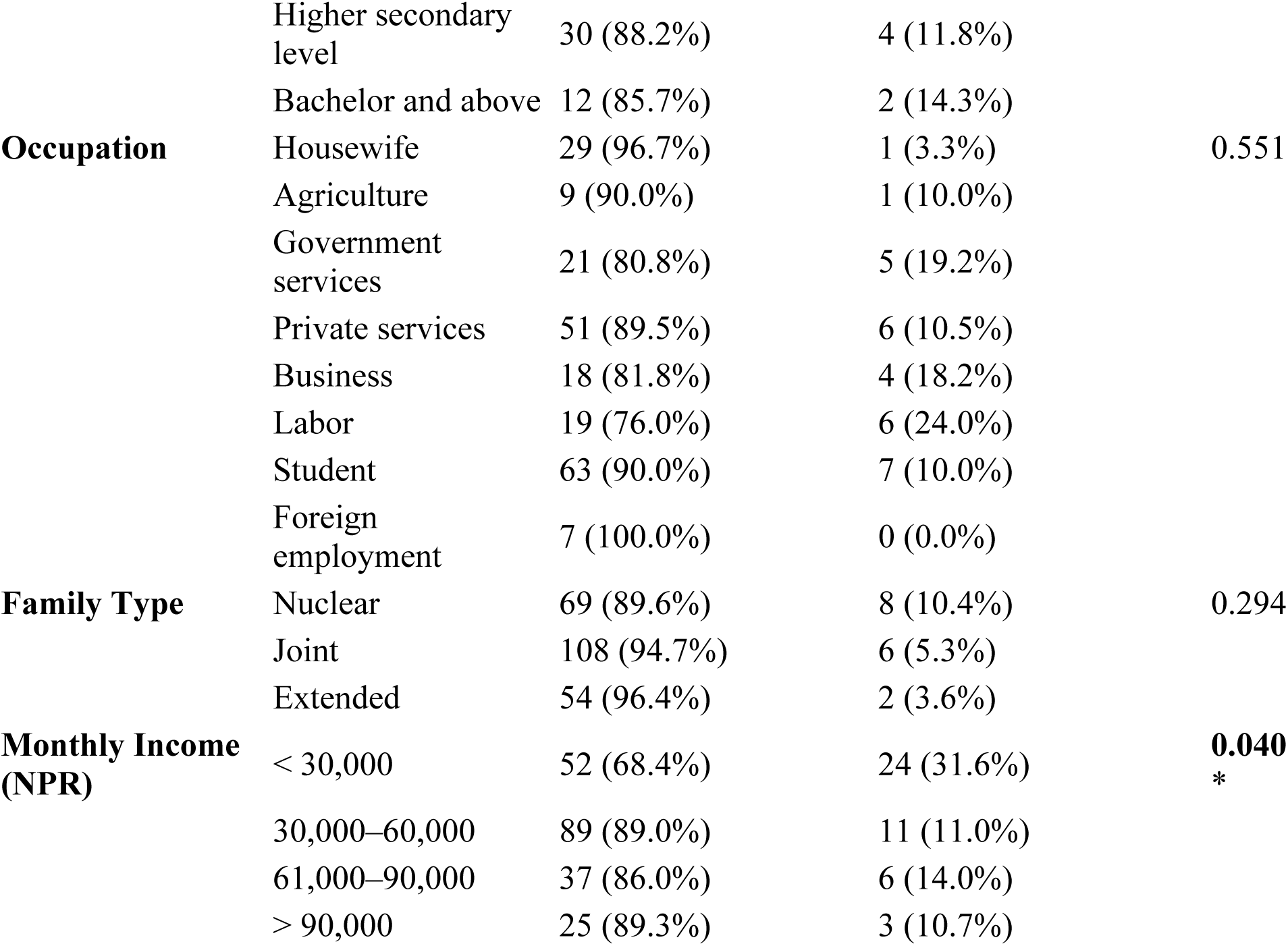
Association between Socio-Demographic Characteristics and Perceived adequacy of the explanation of menstruation.

There was no statistically significant association between age and perceived adequacy (p = 0.054). However, the perception of adequacy was slightly lower among women aged ≥31 years (75.0%) compared to younger age groups, where over 90% reported receiving adequate explanations.

No significant differences were found based on religion (p = 0.439). Adequacy perception was highest among Muslim women (100%) and lowest among Christian women (66.7%), but the differences were not statistically significant.

Similarly, ethnicity was not significantly associated with perceived adequacy (p = 0.288). While Brahmin and Muslim women reported 96.8% and 100% adequacy respectively, slightly lower adequacy was seen among Madhesi (84.6%) and Dalit (85.7%) women.

Marital status did not show a significant relationship with perceived adequacy (p = 0.206). The majority of both married (94.2%) and unmarried (93.7%) women reported adequate explanations, whereas separated women showed a notable contrast, with only 50% reporting adequacy.

Although education status was not significantly associated with perceived adequacy (p = 0.558), a trend was observed: women with lower education levels, particularly those unable to read and write, had the lowest perception of adequacy (55.6%), while those with secondary or higher education generally reported better adequacy (over 85%).

Occupation was not significantly related to adequacy perception (p = 0.551). Housewives and women in foreign employment reported the highest levels of adequacy (96.7% and 100%, respectively), whereas those in labor (76.0%) and business (81.8%) reported lower adequacy.

There was no significant association between family type and perceived adequacy (p = 0.294). Extended families reported the highest adequacy (96.4%), followed by joint (94.7%) and nuclear families (89.6%).

However, a statistically significant association was found between monthly income and perceived adequacy (p = 0.04). Women with a monthly income less than 30,000 NPR had the lowest adequacy perception (68.4%), while those with higher income levels reported better adequacy, reaching up to 89.3% among those earning more than 90,000 NPR per month.

Table 6 presents the association between handwashing practices during menstruation and various socio-demographic characteristics of the respondents. Overall, 31.8% of participants reported using water only, while 68.2% used soap and water for hand hygiene during menstruation.

**Table 6.**
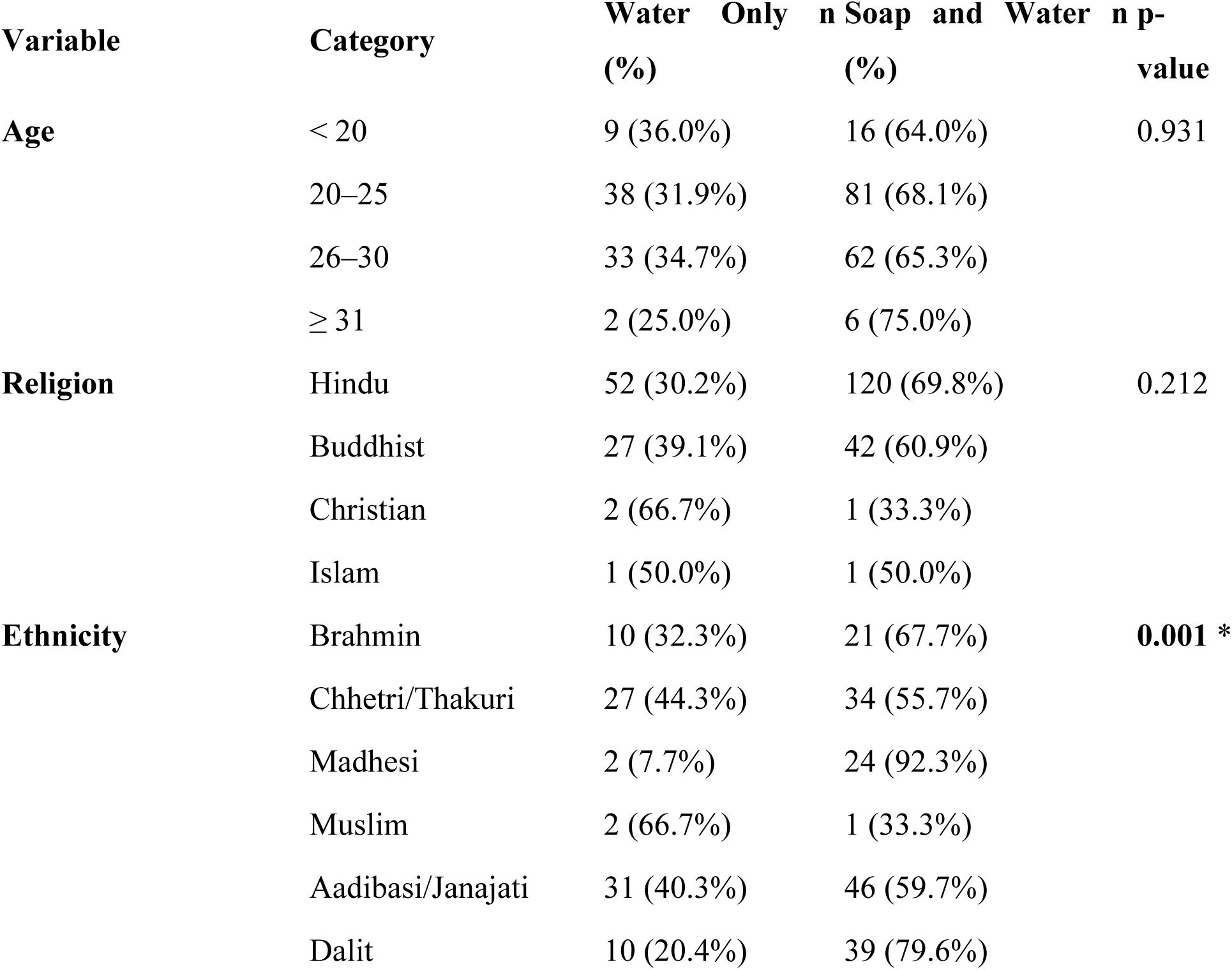
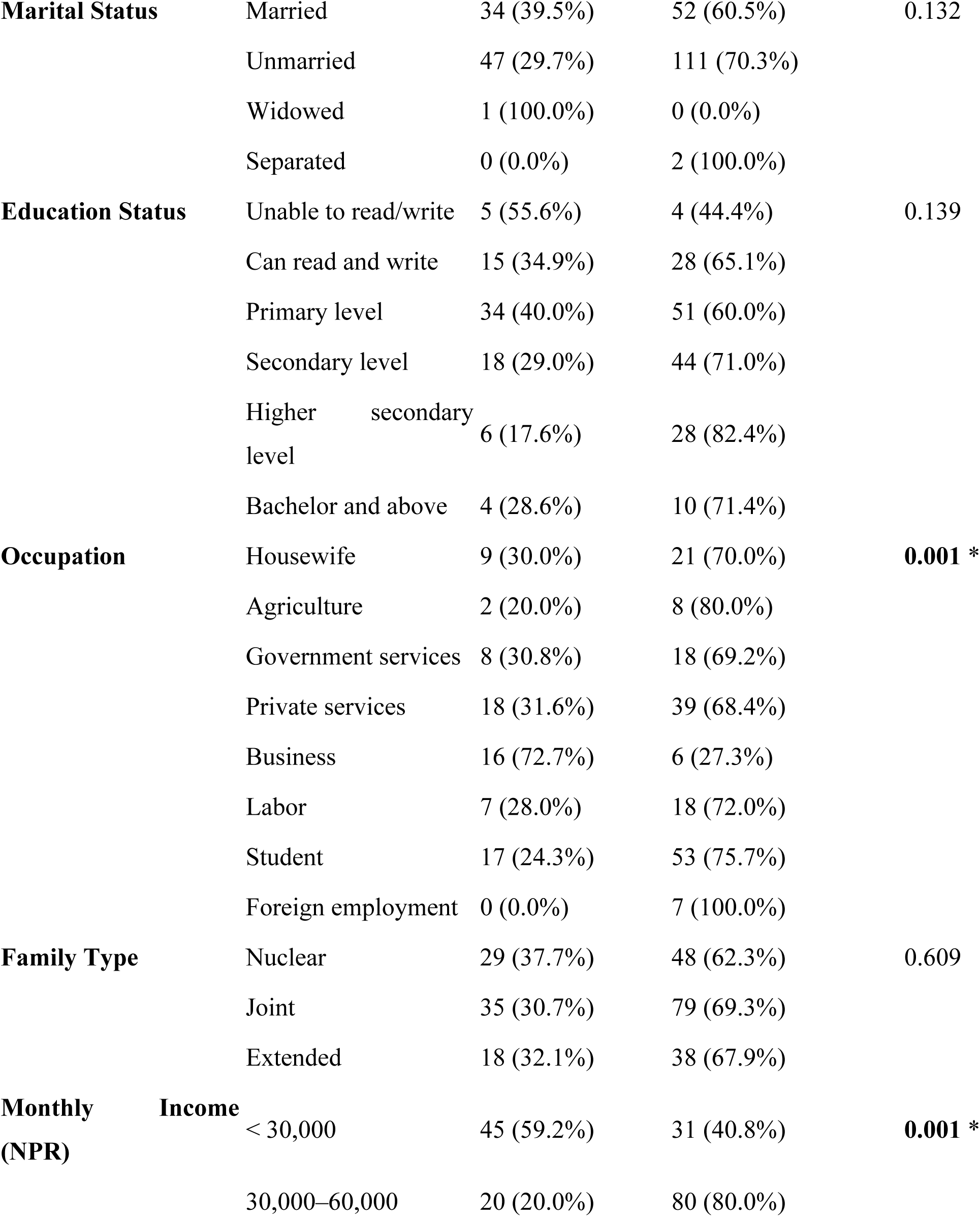
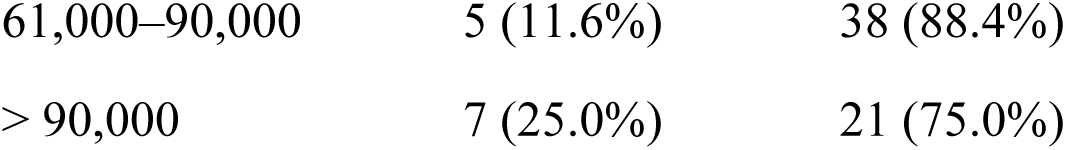
Association between Socio-Demographic Characteristics and Hand wash practices followed by respondent.

There was a statistically significant association between handwashing practices and ethnicity (p = 0.001). A higher proportion of Madhesi (92.3%) and Dalit (79.6%) respondents reported using soap and water compared to other ethnic groups. Similarly, a significant association was observed with occupation (p = 0.001). All respondents in foreign employment and the majority of students (75.7%) reported using soap and water, whereas a higher proportion of businesswomen (72.7%) used only water.

Monthly household income was also significantly associated with handwashing practices (p = 0.001). Respondents with a monthly income below NPR 30,000 had the lowest soap usage (40.8%), whereas those with an income of NPR 61,000–90,000 reported the highest (88.4%).

No significant associations were found between handwashing practices and other variables such as age (p = 0.931), religion (p = 0.212), marital status (p = 0.132), educational status (p = 0.139), and family type (p = 0.609). However, a trend toward improved hygiene with higher education and income levels was noted

Table 7 shows the association between respondents’ socio-demographic characteristics and their practices regarding the cleaning of external genitals during menstruation. The majority of participants reported cleaning with water only, while a smaller proportion used soap and water.

**Table 7.**
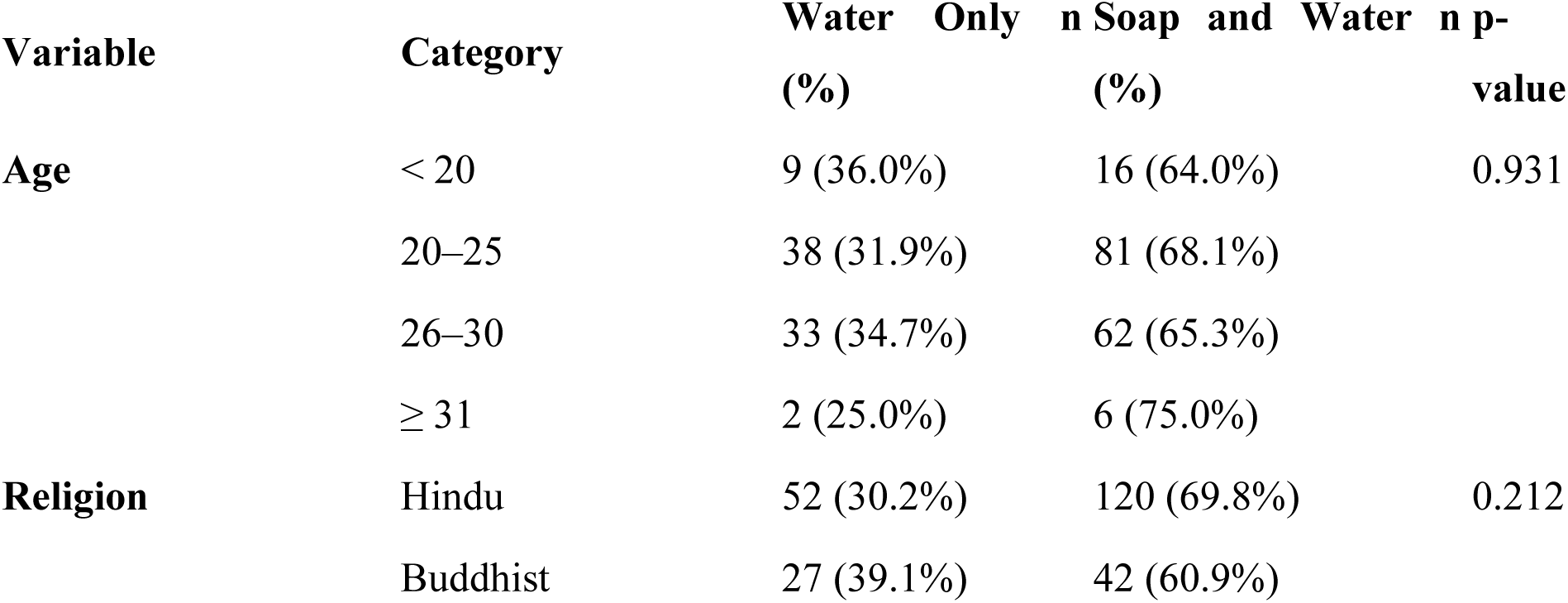
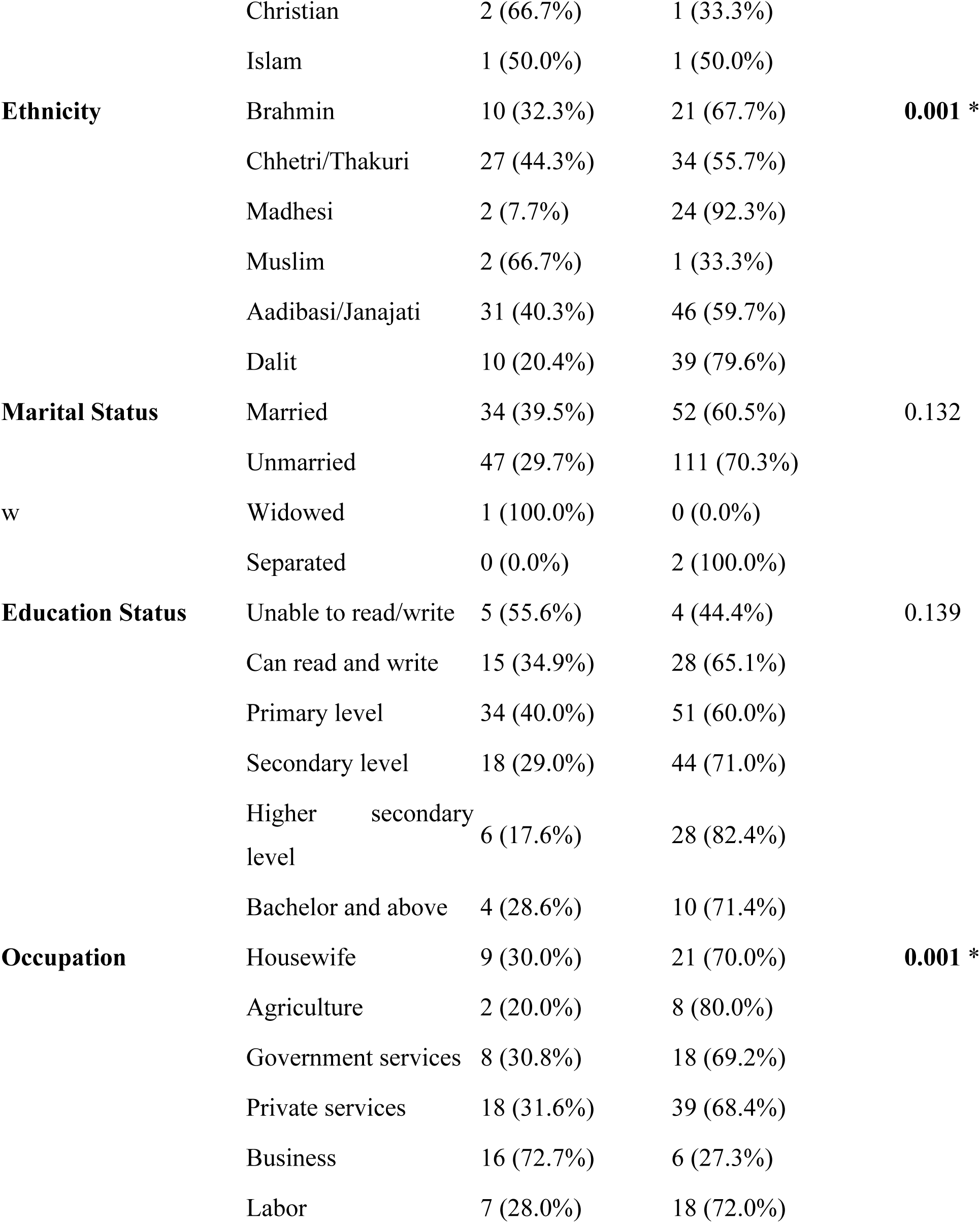
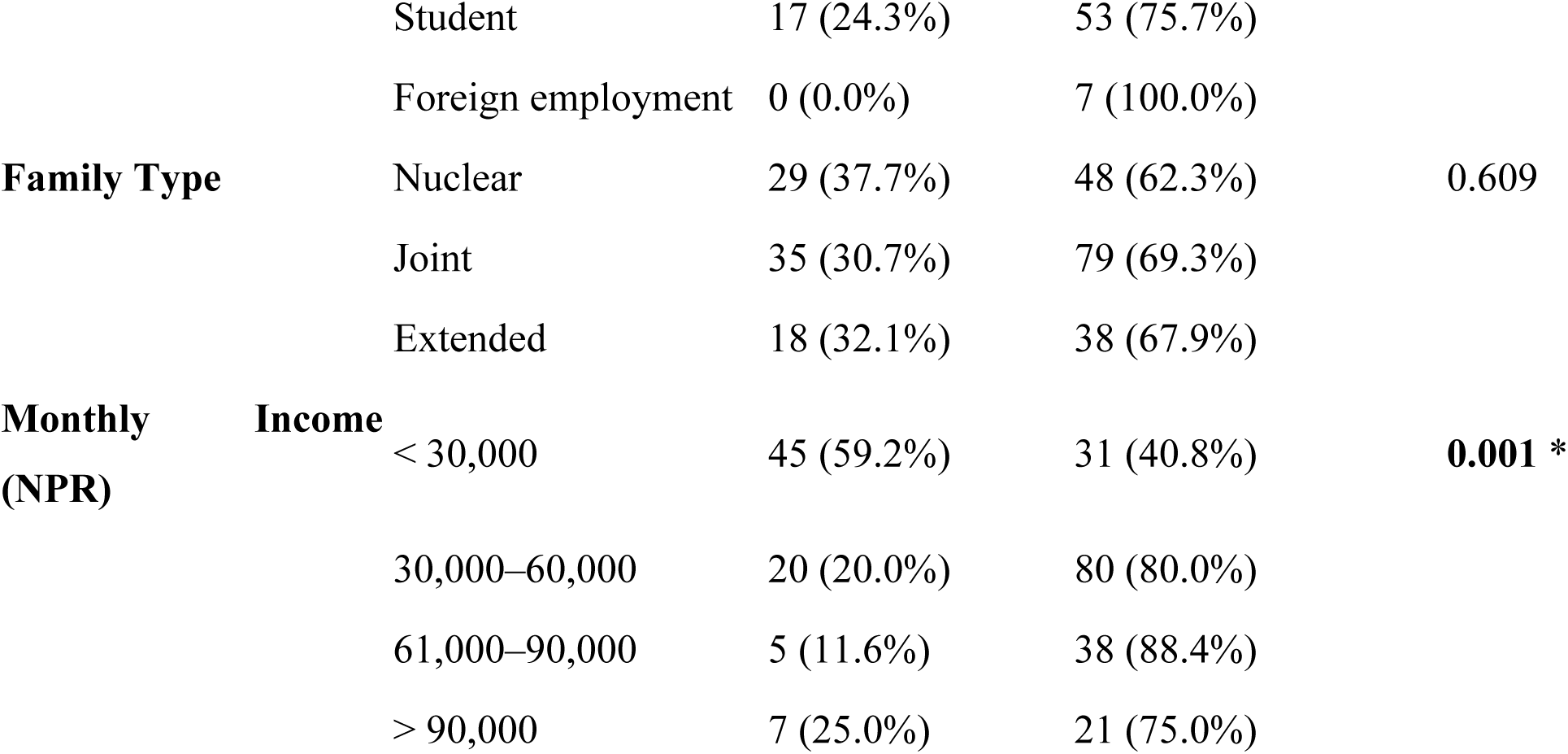
Association between Socio-Demographic Characteristics and Cleaning of external Genitals.

A statistically significant association was observed between family type and cleaning practices (p < 0.001). Among respondents from extended families, 26.8% reported using soap and water compared to only 5.2% in nuclear families and 7.0% in joint families.

No significant associations were observed with age (p = 0.08), religion (p = 0.501), ethnicity (p = 0.161), marital status (p = 0.667), education level (p = 0.141), occupation (p = 0.126), or monthly income (p = 0.207)

This study aimed to identify the factors affecting menstrual hygiene practices among women living in the slum areas of Budanilkantha Municipality, Kathmandu. The findings highlight the influence of socio-demographic variables, cultural taboos, and accessibility to resources on menstrual hygiene practices. In our study, 82.6% of respondents cleaned their external genitals with soap and water, and 66.8% washed their hands with soap and water after changing absorbents. However, the frequency of bathing varied, with only 38.9% reporting daily bathing during menstruation. These practices are essential to prevent infections; however, barriers such as limited access to water, lack of private bathing spaces, and prevailing social norms in slums may hinder hygienic practices. This is in line with the findings from Goddard and Sommer, who emphasized that urban slum populations often face infrastructural limitations that impede menstrual hygiene management (MHM) [6]. The study also revealed that cultural taboos and restrictions were widespread. About 95.1% of participants faced some form of restriction during menstruation, including exclusion from household chores, celebrations, and religious practices. Moreover, 57.1% of respondents reported being considered "impure" during menstruation. These findings align with regional studies which show that menstruation is often surrounded by secrecy and stigma in South Asia [8,13]. However, in a study from urban slums in Karad, India, only 12.6% of adolescent girls used sanitary napkins while the majority relied on reusable cloths [9] These differences may be attributed to varying levels of access to menstrual products, socio-economic status, and education. Despite 65.6% of respondents reporting awareness of menstruation before menarche, the data indicate that knowledge does not always translate into practice. For instance, only 53.8% used sanitary pads, while 42.9% used cloth, which may not always be hygienic. This reflects findings from Deshpande et al., where only 60% of girls used sanitary pads and a majority lacked accurate knowledge of menstruation [10]. The mean age of respondents in this study was 24.8 years, and 53.8% reported using sanitary pads. This finding is comparable to the results from a study conducted in Kathmandu slums, where 65.6% of participants reported using sanitary pads and 96.1% practiced perineal cleaning during menstruation [11]. A significant association was observed between monthly income and awareness of menstruation before its onset (p = 0.002), as well as the perceived adequacy of information received (p = 0.04). This supports the findings from Habtegiorgis et al., who found that maternal education and household income significantly influenced good menstrual hygiene practices in Ethiopia [12]. Similarly, Bhusal et al. also observed a strong association between parental education and menstrual hygiene practices in Nepal [16]. The influence of ethnicity, occupation, and income on handwashing and cleaning practices was statistically significant (p < 0.05). These findings are supported by Chauhan et al., who reported that education level and socio-economic status are predictive of sanitary product usage and hygiene behavior [14].

This finding is consistent with global evidence highlighting that slum dwellers face compounded challenges related to poor sanitation, unsafe housing, and inadequate access to services, which impact menstrual hygiene practices and overall health [15].

While this study provides a comprehensive overview of menstrual hygiene practices in urban slums, further attention is warranted on enabling environmental factors and policy-level gaps. The findings indicate that although a majority of women used soap and water for hygiene, only 38.9% bathed daily during menstruation, and a substantial number (18.6%) did not bathe at all. These figures reflect broader WASH (Water, Sanitation, and Hygiene) infrastructure deficits in slum communities. According to WHO and UNICEF, the availability of private, safe, and clean spaces for bathing is essential for menstrual hygiene management (MHM), yet remains lacking in informal settlements globally [17]. Studies from sub-Saharan Africa and South Asia have emphasized how shared or unsafe toilets and bathing areas prevent women from practicing MHM effectively, often leading to embarrassment and avoidance of hygiene routines [18–19]. Another critical area insufficiently addressed in prior literature but relevant here is the psychosocial impact of menstrual stigma. Nearly 82% of women in this study reported facing social problems, and more than half felt perceived as "impure." While many studies quantify stigma, recent work by Tellier et al. (2021) argues that menstrual stigma contributes to long-term mental health consequences, including shame, low self-esteem, and social isolation, particularly among marginalized women and girls [20]. These effects are often compounded in slum settings where the intersection of poverty and gender inequity limits resilience and access to mental health support. Disposal practices of menstrual products also deserve greater attention. Although 61.9% of women used dustbins, 38.1% reported burning used absorbents. Open burning of sanitary waste, especially in dense urban settings, has environmental and health implications. A review by Van Eijk et al. (2019) highlighted that improper disposal methods are common in LMICs due to limited access to incinerators or sanitary disposal infrastructure [21]. The lack of a safe and culturally appropriate waste management system can discourage the use of disposable pads, further reinforcing cloth usage and suboptimal hygiene practices.

Policy-level attention remains insufficient despite increasing awareness of MHM. While Nepal has made strides by including menstrual health in school curricula and implementing initiatives such as pad distribution programs, these efforts rarely reach out-of-school women or those in informal settlements [22]. A gender-equity-focused approach must extend beyond adolescent girls in schools to include community-based interventions for adult women, especially in socioeconomically disadvantaged urban populations. A recent policy analysis by Mahon et al. emphasized that integrating MHM with broader sexual and reproductive health programs and urban planning is essential for sustainability [23] .

The role of digital media and technology also represents an emerging opportunity. Only 11.6% of respondents in this study reported social media as a source of menstrual knowledge. However, recent findings suggest that mobile-based interventions and health apps can play a role in reaching marginalized groups with culturally sensitive MHM education [24]. Scaling up these tools could improve health literacy and challenge stigma where traditional communication channels fall short. Overall, this study reinforces the need for comprehensive MHM interventions that include awareness programs, improvement in WASH infrastructure, and efforts to challenge harmful social norms. The findings emphasize that MHM is not only a health issue but also a matter of dignity, equity, and rights for women and girls living in marginalized communities.

## Conclusions and Recommendations

A complex interplay of socio-demographic factors, cultural restrictions, and access to resources shapes menstrual hygiene practices among women in the slum areas of Budanilkantha Municipality. Despite moderate awareness and adoption of hygienic practices such as using sanitary pads and washing with soap and water, significant gaps remain—especially in bathing frequency and safe disposal of menstrual products. The pervasive cultural stigma and restrictions faced by nearly all women underscore the urgent need for interventions that address not only knowledge and infrastructure but also social norms and taboos. Enhancing menstrual health in such marginalized urban settings requires comprehensive strategies combining education, improved WASH facilities, stigma reduction, and inclusive policies that empower women and promote dignity and equity. Programs promoting menstrual health should integrate educational, infrastructural, and social support interventions to address multifactorial barriers faced by slum women

## Data Availability

All relevant data are within the manuscript and its Supporting Information files

## Declaration

### Consent for publication

Not Applicable

### Availability of data and materials

All relevant data are included within the manuscript and its supporting information files.

## Acknowledgements

The authors thank all study participants and local authorities for their cooperation

## Authors’ contributions

BLN and AA conceptualized the study. BLN, AA, MP, and PA were involved in data curation. BLN, AA, and PA performed formal analysis. The methodology was developed by BLN, AA, and PA. Supervision was provided by PA. Validation was conducted by BLN, AA, MP, and PA. The original draft was written by BLN, AA, MP, and PA, with review and editing by BLN, AA, and PA. All authors read and approved the final manuscript.

